# Effects of PM2.5 Air Pollutants on Cardiovascular Health in Ohio: A Secondary Data Analysis

**DOI:** 10.1101/2022.10.19.22281274

**Authors:** Peyton G. Finkle, Jahasia W. Gillespie, Brian J. Piper

**Affiliations:** Geisinger Commonwealth School of Medicine, Scranton, PA 18509; Center of Pharmacy Innovation and Outcomes, Geisinger Precision Health Center, Forty Fort, PA 18704

## Abstract

**Background:** Multiple adverse health outcomes have been linked to air pollution, particularly exposure to particulate matter (PM_2.5_).

**Justification:** This research will help to identify the role that socioeconomic and geographical factors have in increased cardiovascular disease and cardiac events due to elevated exposure amounts of PM_2.5_. Minorities in the United States are more likely to have or die from cardiovascular disease and also are the majority of the occupants in low-income communities with high levels of PM_2.5_.

**Methods:** PM_2.5_ pollutant concentrations were measured from 2016 to 2018 and collected by air quality monitoring equipment in Ohio. The cardiovascular disease data on adult residents of Ohio from 2016-2018 was from the National Vital Statistics System (NVSS) and the Centers for Medicare and Medicaid Services Medicare Provider Analysis and Review (MEDPAR). R programming was used to find associations between cardiovascular disease and PM_2.5_ in Ohio. Contingency tables compared the consumption of PM_2.5_ and the rate of heart disease mortality. Heat maps indicated areas where heart disease mortality and PM_2.5_ concentrations were highest.

**Results:** The average PM_2.5_ concentration in Ohio was determined to be 7.9 μG/m^3^. The highest PM_2.5_ concentrations were found in Montgomery County, Ohio and the lowest were recorded in Allen County, Ohio. With increased population density, there was also an increase with PM_2.5_ concentrations (P<0.01). The association between cardiovascular disease mortality and PM_2.5_ rates in these counties was nonsignificant (P>0.01). It was also found that 31.8% of counties in Ohio had air monitoring while the remaining 68.2% did not have any form of it.

**Conclusion:** Knowing the risk of PM_2.5_ levels in any neighborhood could help community members and leaders pick a more appropriate course of action. Reducing air pollutants in these areas may in turn reduce cardiovascular disease in that population, leading to better health outcomes and longevity. This research also identified gaps in air monitoring that assists in prioritizing areas that need immediate remediation.

## Introduction

### Background

Particulate matter (PM_2.5_) is a combination of solid particles and liquid droplets of pollutants found in the air with a size of 2.5 micrometers or less in diameter (1). The first entry point and primary deposition of these particles is through the lungs (2). Because of their size, smaller particles can travel deeper into the lungs causing respiratory damage. Long-term exposure to PM_2.5_ has increased, with average concentrations ranging from approximately 5 to 30 μg/m^3^ (3). Once absorbed into the bloodstream, they can reach anywhere in the body, causing premature aging of blood vessels and contributing to a rapid buildup of calcium in the coronary artery, causing cardiovascular disease (4,5).

Recent studies have connected elevated PM_2.5_ levels with cardiopulmonary disorders and impairments, the initiation and progression of diabetes mellitus, and poor birth outcomes (2). Fractions of PM_2.5_ are retained in the lungs and account for roughly 96% of particles observed (1). Another study outlined the association of PM_2.5_ exposure and robust lung inflammation causing vascular remodeling, as well as exacerbated transition from left ventricular failure to right ventricular hypertrophy (6). Findings suggested that air quality was a critical factor in overall heart failure progression by regulating lung inflammation and remodeling (6). Two particularly vulnerable groups are women and children. PM_2.5_ has been linked to neonatal and post neonatal mortality, premature births, and acute respiratory infections (6,7).

According to the CDC, cardiovascular disease is the leading cause of death in the United States (U.S.) (4,8). Approximately 15 million Americans were diagnosed with asthma, with around 5,000 dying annually (9). Other studies have identified air pollution as a large environmental contributor to health outcomes not only in the U.S., but globally as well (10). Air pollution has been repeatedly associated with higher cardiovascular morbidity and mortality (10). Numerous studies have linked elevated exposure of PM_2.5_ to adverse health effects such as cardiovascular disease and birth complications (11,12). Many investigations associate the relationship between the acquisition and the mechanisms in which PM_2.5_ leads to adverse effects in the body (12,13). However, there are not many studies relating the geographic locations of landfills, manufacturing plants, and other air pollutant sources near low-income communities to the increased rate of acquisition of cardiovascular disease and asthma, resulting from elevated PM_2.5_ (14).

Our primary objective was to determine if socioeconomic or geographic factors contribute to the acquisition of asthma and cardiovascular disease in low-income communities. Through the use of spatio-temporal models, PM_2.5_ can be compared with socioeconomic position and racial composition (15). Low income, minority communities, including African Americans, Asians, and Hispanics are more likely to develop cardiovascular disease and other respiratory related diseases due to high PM_2.5_ level exposure (15,16). Despite the wide demographic within low-income communities, African Americans have exhibited higher rates of cardiovascular disease and asthma than any other ethnic background. This fact indicates that there may be factors outside of geography and socioeconomic factors that are contributing to adverse health effects resulting from PM_2.5_ (17,18). This research may aid in the development of future communities and give birth to new laws and regulations on the placement of residential areas in areas emitting high concentrations of pollutants.

A secondary objective was to compare Franklin and Vinton County, which were differentiated by population. Franklin County held the highest population per county in the state at 1,323,807 residents, whereas Vinton County held the lowest population per county in Ohio at 12,800 residents per the 2020 census (19). Franklin County is located in the northeastern region of the state where there is a high population density. Vinton County, however, is in the southeastern region of the state, where the population density is lower. This research examined if infrastructure growth of these counties leading to increased PM_2.5_ trends increases with their respective populations.

The tertiary objective was to examine the completeness of the air monitoring infrastructure and capabilities across the counties in Ohio.

## Methods

### Participants

The study population was the adult residents of Ohio whose health data was collected and reported. The inclusion criterion for this population was age (>18 years), and participants also had to be diagnosed with cardiovascular disease or experience cardiovascular disease related death (<75 years). The exclusion criteria were obesity, and the genetic acquisition of cardiovascular disease as previous research has linked cardiovascular disease to involuntary weight gain (20), genes, and specific DNA sequence variants (21). This study was approved exempt by the Geisinger Commonwealth School of Medicine IRB.

### Data Exploration & Inferential Statistics

The goal of this research was to find associations between cardiovascular disease with the daily mean intake of PM_2.5_ in Ohio. Using descriptive analysis in the form of contingency tables and heat maps aided to visualize the data collected and draw conclusions. Heat maps were used as a spatial representation of the magnitude of PM_2.5_ and cardiovascular disease development and mortality rates per county. R version 4.1.2 and the Integrated development environment (IDE) R studio version 2022.02.0+443 for macOS was used to perform calculations and generate graphs.

## Results

The relationship between cardiovascular disease mortality and the daily mean PM_2.5_ concentration was illustrated (Table 1). Among the 88 counties in Ohio, the median population density was 4 according to the rural-urban continuum code updated decennially (22). The lowest population density coefficient was 8, indicating a completely rural or less than 2,500 urban population that is adjacent to a metro area (Table 1). The counties in Ohio that fell into this category were Monroe and Vinton County. The highest population density was 1, including counties considered to be in metro areas of 1 million population or more. Of these were 20 counties in Ohio, including Franklin County, which is the county where Ohio’s capital Columbus resides.

**Table 1.**
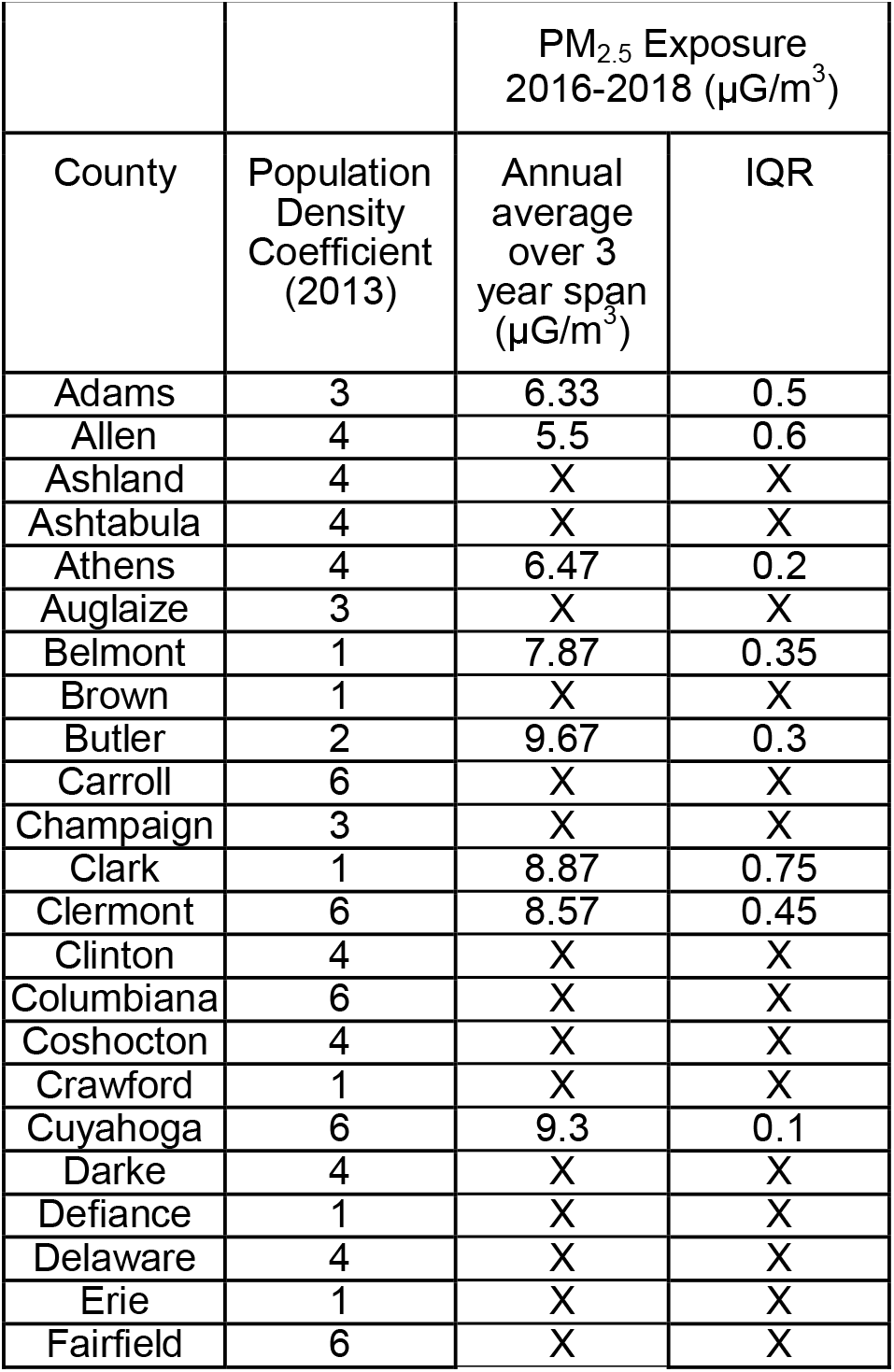
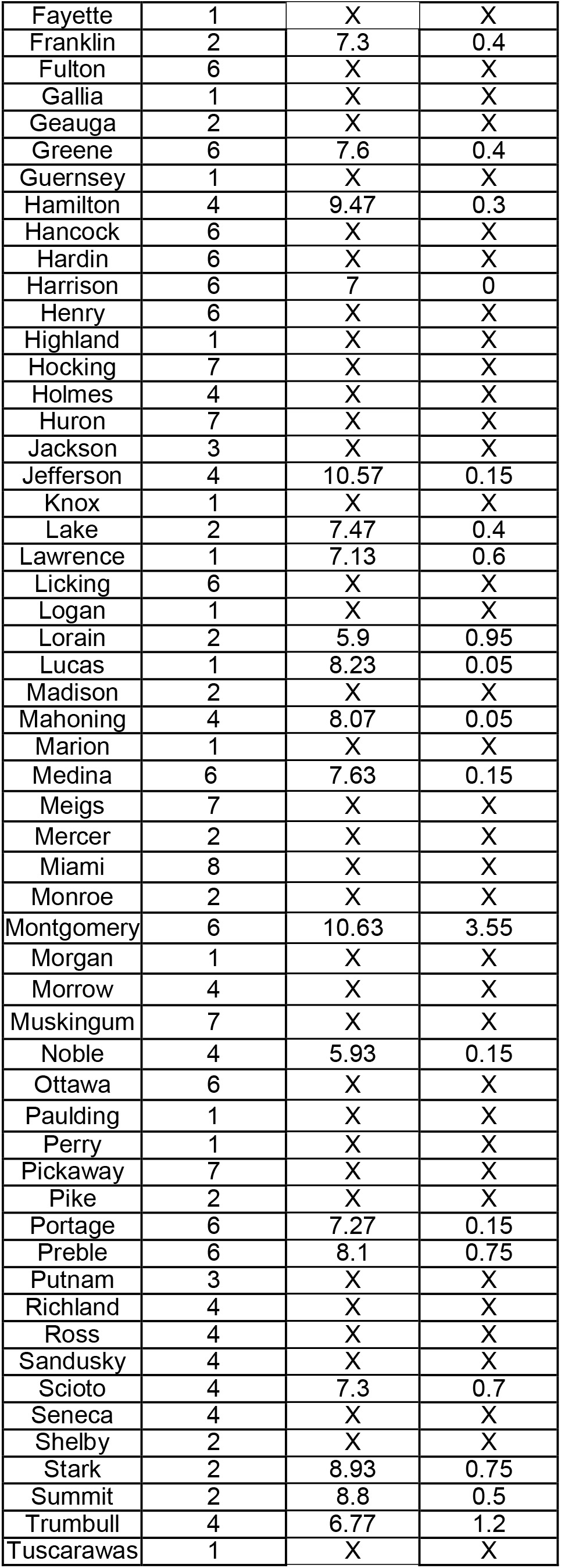
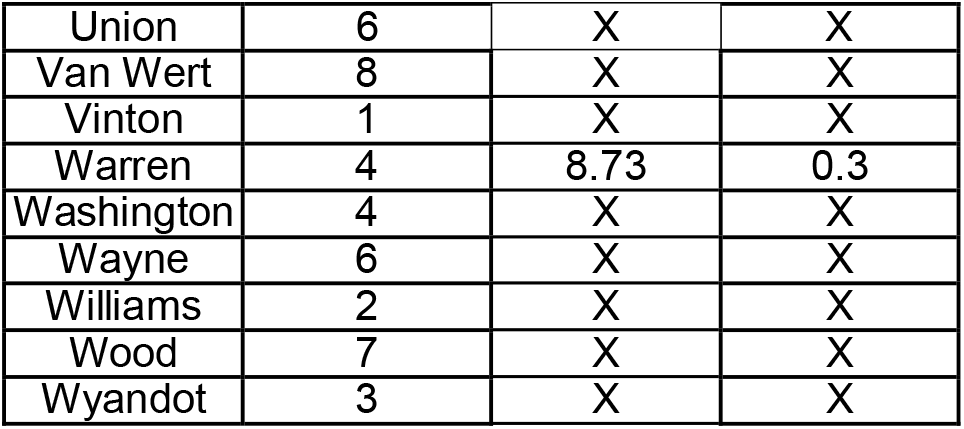
This table depicts counties and their respective population density coefficients. Triannual averaged PM_2.5_ concentrations are available for the counties equipped with AQI monitoring data. Interquartile ranges (IQR) were also provided for these PM_2.5_ concentrations.

Summarizing PM_2.5_ concentrations from 2016-2018 in Ohio, the average concentration was 7.9 μG/m^3^, which is 41.2 percent lower than the current recommended amount reported by the EPA, 12 μG/m^3^ (23). To examine health effects of exposures in these counties, cardiovascular disease mortality rates for 2016-2018 were compared to PM_2.5_ concentrations. The association between cardiovascular disease mortality and PM_2.5_ rates in these counties was nonsignificant (P>0.01).

Three heat maps were constructed showing various magnitudes of PM_2.5_ concentrations and cardiovascular disease complications within the counties (Figure 1). Levels varied between counties, with the highest PM_2.5_ concentrations in Montgomery County (10.6 μG/m^3^), and the lowest recorded in Allen County (5.5 μG/m^3^) (Figure 1). The interquartile range (IQR) of PM_2.5_ averages was 1.72 alluding to consistent PM_2.5_ concentrations between counties. The association between population density and PM_2.5_ was significant, indicating with increased population density there was also an increase with PM_2.5_ concentrations (P<0.01).

**Figure 1.**
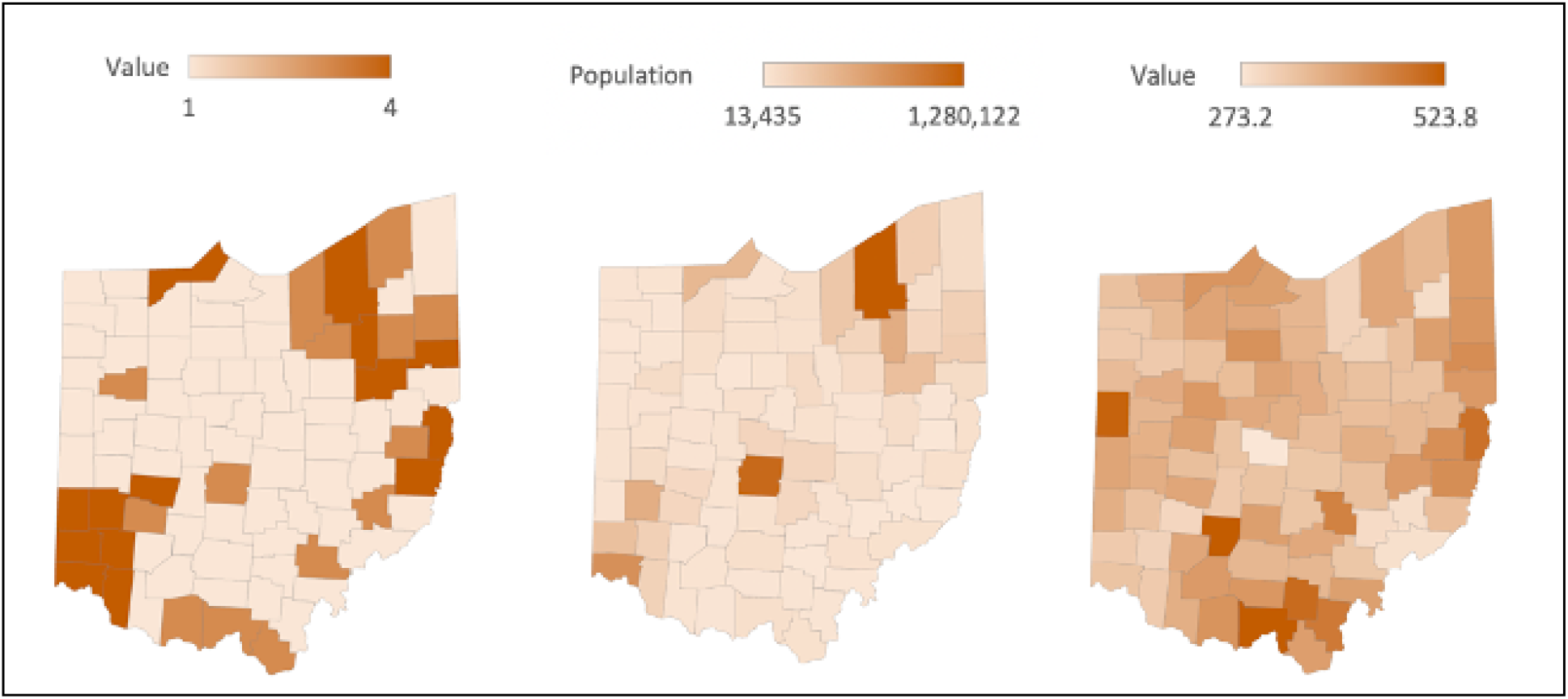
The left panel shows the average PM_2.5_ concentrations by county in Ohio from 2016 to 2018, triannually. A key above the map showcases ratings for each county on a scale from 1-4. Counties with PM_2.5_ values from 2-4 μG/m^3^ were rated a 2, counties with PM2.5 values between 5-7 μG/m^3^ were rated a 3, and counties with PM2.5 values 8-10 μG/m^3^ were rated a 4. The middle panel shows the average population density in each county in Ohio in 2013. The right panel shows the cardiovascular disease mortality per county in Ohio averaged between 2016 and 2018.

To address the completeness of air quality in the state of Ohio, 31.8% of counties in Ohio had air monitoring while the remaining 68.2% did not. More than two-thirds of the state has no form of air quality monitoring at all, meaning the “average” concentrations in Ohio are based on less than one-third of the state’s readings.

## Discussion

Our main objective in this study was to assess the association between PM_2.5_ exposure and incidence of adverse health outcomes across counties in Ohio from 2016-2018. The average concentration of PM_2.5_ was determined to be 7.9 μG/m^3^, which is 41.2 percent lower than the current recommended amount reported by the EPA, which is 12 μG/m^3^(23). Although lower than the EPA standard, these concentrations still prove to be unhealthy for human consumption on a daily basis (2).

Our secondary objective was to compare Franklin and Vinton counties due to their variance in population. The association between population density and PM_2.5_ was significant (P<0.01). The association between cardiovascular disease mortality and PM_2.5_ rates in these counties was nonsignificant (P>0.01).

The last objective of this study was to examine the completeness of the air monitoring capabilities across the counties in Ohio. The number of counties with air quality monitoring was scarce, with 31.8% of counties in Ohio having adequate air monitoring. Therefore, more than two-thirds of the state had no form of air quality monitoring at all, basing the “average” concentrations in Ohio on less than one-third of the state’s actual readings. The lack of data due to sparse equipment availability for lower income, rural, and minority communities limited overall data output for the state of Ohio as a whole. In the future, infrastructure should be created in these areas to take a fair approach to air quality monitoring. Without data to assess the quality of the air, conclusions cannot be made as to what role it is playing in the health of the community overall.

## Conclusion

Despite the efforts in recent years to reduce vehicle emissions and other contributors to poor air quality, the current efforts in Ohio seem to be inadequate, especially in more rural counties (24).

Several studies done in the US have indicated that asthma is more common in urban settings and in individuals who are African American, Hispanic, male, or live in lower socioeconomic areas (15,16,17,18). This furthers the need for research inclusive of socioeconomic or geographic factors to find solutions to benefit the communities who are most at risk. This research can serve as a basis for the creation of prevention strategies and mitigation techniques. For decades, there has been a call for environmental justice within the US. Residing in a low-income community is associated with physical and mental health risks (14). Hopefully with the information derived from this study areas of priority are highlighted to create a more sustainable, equitable future for Ohio (25).

## Data Availability

All data produced in the present work are contained in the manuscript.

## Acknowledgements

Theo Light, PhD, Oritsetosan S. Emmanuel, MBS, and Timerra G. Hardy, MBS, provided technical assistance.

## Notes

### Competing Interest Statement

The authors have declared no competing interest.

### Funding Statement

This study did not receive any funding.

### Author Declarations

This study was approved exempt by the Geisinger Commonwealth School of Medicine IRB.

